# OBESITY MAY HAMPER SARS-CoV-2 VACCINE IMMUNOGENICITY

**DOI:** 10.1101/2021.02.24.21251664

**Authors:** Raul Pellini, Aldo Venuti, Fulvia Pimpinelli, Elva Abril, Giovanni Blandino, Flaminia Campo, Laura Conti, Armando De Virgilio, Federico De Marco, Enea Gino Di Domenico, Ornella Di Bella, Simona Di Martino, Fabrizio Ensoli, Diana Giannarelli, Chiara Mandoj, Valentina Manciocco, Paolo Marchesi, Francesco Mazzola, Silvia Moretto, Gerardo Petruzzi, Fabrizio Petrone, Barbara Pichi, Martina Pontone, Jacopo Zocchi, Antonello Vidiri, Branka Vujovic, Giulia Piaggio, Aldo Morrone, Gennaro Ciliberto

**Author notes:** Corresponding author: Aldo Venuti, MD, PhD, IRCCS “Regina Elena” National Cancer Institute, Via Elio Chianesi 53, 00144, Rome – Italy.

## Abstract

**Background:** The first goal of the study was to analyse the antibody titre 7 days after the second dose of BNT162b2 vaccine in a group of 248 healthcare workers (HCW). The second goal was to analyse how the antibody titre changes in correlation with age, gender and BMI.

**Methods:** Participants were assigned to receive the priming dose at baseline and booster dose at day 21. Blood and nasopharyngeal swabs were collected at baseline and 7 days after second dose of vaccine.

**Findings:** 248 HWCs were analysed, 158 women (63.7%) and 90 men (36.3%). After the second dose of BNT162b2 vaccine, 99.5% of participants developed a humoral immune response.

The geometric mean concentration of antibodies among the vaccinated subjects after booster dose (285.9 AU/mL 95% CI: 249.5-327.7); was higher than that of human convalescent sera (39.4 AU/mL, 95% CI: 33.1-46.9), with p<0.0001. The antibody titre was found to be higher in young and female participants. A strong correlation of BMI classes with antibody titres was noticed: humoral response was more efficient in the group with under- and normal-weight vs the group with pre- and obesity participants (p<0.0001 at T1).

**Interpretation:** These findings imply that females, lean and young people have an increased capacity to mount humoral immune responses compared to males, overweight and the older population. Although further studies are needed, this data may have important implications for the development of vaccination strategies for COVID-19, particularly in obese people.

**Funding:** None

## INTRODUCTION

Since the first cases of COVID-19 were described in December, a health emergency with major social and economic disruptions has spread worldwide.

The World Health Organization, on the 11th of March 2020, announced the severe acute respiratory syndrome coronavirus 2 (SARS-CoV-2) outbreak a pandemic.

Although rigorously applied, control measures such as the use of masks, physical distancing and contact tracing, helped to limit viral transmission; however no substantial benefits were noted, and it soon became clear that vaccines represented the main viable road to get out of the pandemic.

Since the genetic sequence of SARS-CoV-2 on January 11, 2020, scientists and biopharmaceutical manufacturers focused their research on developing a vaccine.

Currently, there are over 238 vaccine candidates being developed against COVID-19, with 63 at various stages of human clinical trial test(1).

A large clinical trial phase 2/3 with 44,000 people showed that a two-dose regimen of the vaccine BNT162b2, developed by BioNTech and Pfizer, has 95% efficacy in preventing symptomatic COVID-19. The same study showed that safety over a median of 2 months was similar to that of other viral vaccines(2). As a consequence of these results, on December 11th, 2020, the U.S. Food and Drug Administration authorized vaccine BNT162b2 for emergency use. This was soon followed by the European Medicines Agency on 21 December 2020.

Nonetheless, the efficacy of protection from infection of BNT162b2 vaccine has not been established. To overcome this limit, antibody titre can be used to predict protection against SARS-CoV-2, as already done for many viruses in humans and for SARS-CoV-2 in animal challenges(3, 4).

Two studies by BioNTech/ Pfizer’s presented immunogenicity data: preliminary results are encouraging, but antibody responses with a double 30 micrograms regimen is reported only in 22 patients(5, 6).

Krammer et al, described the antibody responses in 109 individuals with and without documented pre-existing SARS-CoV-2 immunity (seronegative: 68, seropositive: 41). They advised that a single dose of mRNA vaccine elicits very rapid immune responses in seropositive individuals with post-vaccine antibody titres that are comparable to or exceed titres found in naïve individuals who received two vaccinations(7). More data is certainly needed to assess the efficacy and thus protection against the virus.

In this setting, we report the early experience with BNT162b2 vaccination in a medical population. The first goal of our study was to analyse the antibody titre response 7 days after the second dose of vaccine in a group of 248 healthcare workers (HCW). Our second goal was to analyse how the antibody titre changes in correlation with age, gender and BMI.

## MATERIALS AND METHODS

### Study design and participants

A collaborative team carried out an immunogenicity evaluation among HCWs vaccinated at the Istituti Fisioterapici Ospitalieri (IFO).

The study protocol complied with the tenets of the Helsinki declaration and was approved by the institutional scientific ethics committee (protocol RS1463/21).

All the enrolled participants met the following inclusion criteria: 1) provided written informed consent 2) age between 18-75 years, 3) health workers employed at the Istituti Fisioterapici Ospitalieri (IFO), 4) vaccinated at the Istituti Fisioterapici Ospitalieri (IFO).

Key exclusion criteria included: 1) evidence of current or previous SARS-CoV-2 infection by either anamnesis, serological or microbiological test through nasopharyngeal swab before enrolment, 2) treatment with immunosuppressive therapy, 3) immunosuppression-associated pathology, and 4) pregnancy.

Human SARS-CoV-2 infection convalescent sera (*n*=59) were drawn from HCW donors (mean age 45) at least 14 days after PCR-confirmed diagnosis and at a time when the participants were asymptomatic. The donors were divided into groups based on symptoms: symptomatic infections (*n*:41); asymptomatic infections (*n*:18).

The manufacturer’s (BioNTech/Pfizer, USA) instructions for storage and administration of vaccine were followed.

The COVID-19 mRNA Vaccine BNT162b2 was stored in an ultra-low temperature freezer at −80°C. The undiluted vaccine was stored for up to 2 hours at temperatures up to 25°C, prior to use.

The mRNA vaccine was administered as a 30 microgram / 0.3ml intramuscular injection into the deltoid muscle on days 1 and 22 of the study.

We used a questionnaire to collect data on the participants’ socio-demographic and health characteristics. Participants were stratified by age, sex and body mass index (BMI).

Participants had a nasopharyngeal swab for SARS-CoV-2 RT-PCR testing (Viracor, Eurofins Clinical Diagnostics, U.S.A.), and were assessed for the presence of SARS-CoV-2–binding antibodies (The LIAISON® SARS-CoV-2 S1/S2 IgG, test Diasorin, Italy) and at baseline and 7 days after BNT162b2 booster dose.

### Assessment of SARS-CoV-2 in nasopharyngeal swab

A nasopharyngeal swab was collected by standard procedures and presence of SARS-CoV-2 was determined by RT-PCR testing (Viracor, Eurofins Clinical Diagnostics, U.S.A.) following the manufacturer’s instruction.

### Assessment of SARS-CoV-2 Binding Antibody

Peripheral venous blood samples of 7-8 mLs were obtained, serum collected and stored at +4 °C.

Quantitative measurement of IgG antibodies against S1/S2 antigens of SARS-CoV-2 was performed with a commercial chemiluminescent immunoassay (The LIAISON® SARS-CoV-2 S1/S2 IgG, test Diasorin, Italy) according to the manufacturer’s instruction.

### Statistical analysis

Log Geometric Mean of AU/mL was reported. To assess differences between groups, Student’s t-test (Bonferroni’s adjusted) was used when comparing between 2 groups and ANOVA when comparing between >2 groups.

Repeated measurement ANOVA was used to assess differences between groups over time.

Age was categorized according to quartiles. Statistical analysis was done using SPSS Statistics software version 21. A *p* < 0.05 was considered statistically significant.

## RESULTS

Two hundred forty-eight HWCs were analysed, 158 women (63.7%) and 90 men (36.3%). Median age was 47 years, (range 23-69). Nasopharyngeal swab test at baseline and 7 days after booster dose did not reveal presence of SARS-CoV-2 in any of the participants.

Results are summarized in table 1. After second dose of BNT162b2, 99.5% of participants developed a humoral immune response respect to the baseline (T0) serum level. Only one participant was non-responder after the second dose.

**TABLE 1:** GMC and 95% CI by age, gender, BMI and hypertension at T0 and T1 (7 days after BNT162b2 booster dose).

| **CHARACTERISTIC** | **NUMBER** | **GMC (95% CI)** | **GMC (95% CI)** |
| --- | --- | --- | --- |
| **SAMPLING** |  | **T0** | **T1** |
| **AGE** |  |  |  |
| <=37 | 62 | 4.0 (3.9-4.1) | 453.45 (363.6-565.5) |
| 37-47 | 63 | 4.3 (4.0-4.6) | 330.91 (253.7-431.6) |
| 47-56 | 64 | 4.1 (3.9-4.4) | 239.77 (182.4-315.1) |
| >56 | 59 | 4.0 (3.9-4.2) | 182.40 (137.8-241.4) |
| **GENDER** |  |  |  |
| FEMALE | 158 | 4.0 (3.9-4.2) | 338.49 (286.3-400.2) |
| MALE | 90 | 4.2 (4.0-4.5) | 212.63 (170.2-265.6) |
| **BMI** |  |  |  |
| UNDER-WEIGHT | 19 | 3.9 (3.7-4.1) | 455.41 (311.5-665.7) |
| NORMAL-WEIGHT | 147 | 4.2 (4.0-4.3) | 325.84 (277.9-382.0) |
| PRE-OBESITY | 56 | 4.0 (3.9-4.2) | 222.40 (168.9-292.9) |
| OBESITY | 26 | 4.2 (3.8-4.7) | 167.05 (90.2-309.3) |
| **HYPERTENSION** |  |  |  |
| NO | 217 | 4.1 (4.0-4.2) | 307.42 (267.4-353.4) |
| YES | 31 | 4.3 (3.9-4.7) | 172.18 (109.0-272.1) |

Note: GMC: geometric mean concentration; CI: confidence interval; BMI: body mass index according to Weir CB and Jan A(10).

Seven days after the booster dose, S1-S2 binding antibody concentration was in the range of 3.8-2460 AU/mL. The antibody geometric mean concentration (AbGMC) among vaccinated subjects (285.9 AU/mL 95% CI: 249.5-327.7) was higher than that of human convalescent sera (39.4 AU/mL, 95% CI: 33.1-46.9), with p<0.0001.

The antibody titre was greater in younger participants compared to older participants with statistically significant differences: <=37 vs 47-56 p=0.005, <=37 vs >56 p<0.0001, 37-47 vs >56 p=0.01. AbGMC of single participants among different age classes is reported in figure 1.

**Figure 1:**
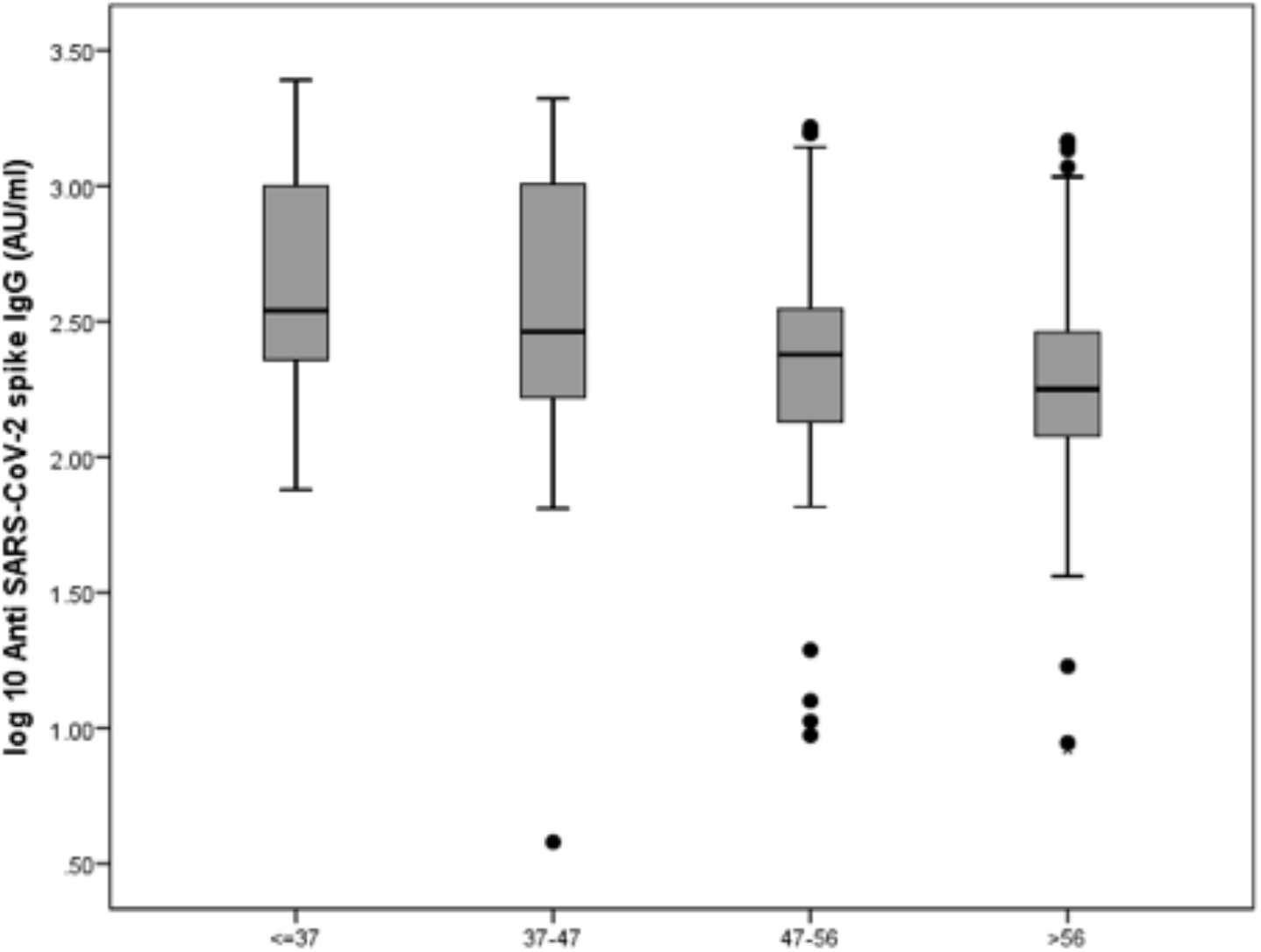
Levels of anti SARS-CoV-2 spike IgG by age classes. Serum was collected from participant’s antibody 7 days after booster dose. Antibody levels were expressed as log10 of concentration in Arbitrary Unit (AU). Age was categorized according to quartiles.

Antibody responses of greater magnitude was shown more frequently in women than in men; this difference (338.5 AU/mL vs 212.6 AU/mL) was statistically significant with p=0.001.

AbGMC of single participants among BMI classes is showed in figure 2. A correlation between BMI classes and antibody titres was noticed with p=0.02. Interestingly, humoral response was more efficient in under- and normal-weight group vs pre-obesity and obesity group (p<0.0001). This association was confirmed after adjusting for age (p=0.003).

**Figure 2:**
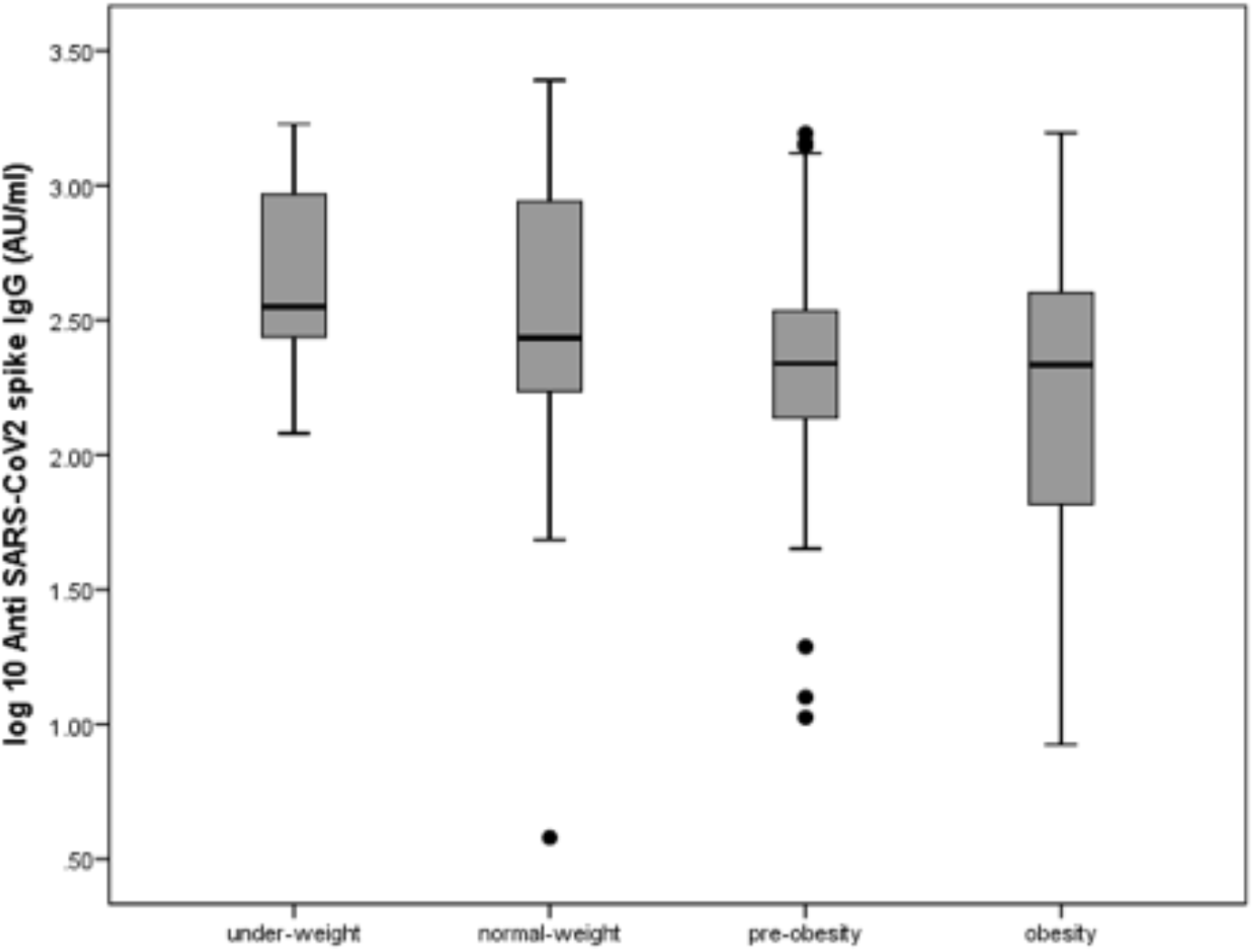
Levels of anti SARS-CoV-2 spike IgG antibody by BMI classes. Serum was collected from participants 7 days after booster dose. Antibody levels were expressed as log10 of concentration in Arbitrary Unit (AU). Body mass index (BMI) classes were categorized according to Weir CB and Jan A(10).

Finally, hypertension was associated with lower AbGMC titre with p=0.006. However, after age matching this data there was no statistical significance found (p=0.22).

## DISCUSSION

During the last year we have witnessed a remarkable effort of researchers and the pharmaceutical industry in the development of a vaccine against SARS-CoV-2.

With this paper, we present an independent study on antibody titre against S1/S2 SARS-CoV-2 in HCWs 7 days after the second dose of BNT162b2: almost 100% of participants demonstrated antigen-specific humoral response respect to baseline level and no one showed positive nasopharyngeal test during the study.Although the role of neutralizing antibodies to SARS-CoV-2 is under investigation, measurement of serum neutralizing activity has been demonstrated to correlate with protection for other respiratory viruses, such as influenza(3) or respiratory syncytial virus(8) and is commonly accepted to be a functional biomarker of in vivo disease protection(9). In this study we utilized a chemiluminescent immunoassay that detect S1/S2 specific antibodies but it was not specifically designed to detect neutralizing antibodies. However, the manufacturer indicates that 80 AU/mL has a correlation of 100% with a 1:160 titre of a Plaque reduction neutralization test (PRNT90). Thus, we can assume that at least a substantial proportion (93.2%) of enrolled population showing >80AU/ml should have developed neutralizing antibodies.

Our study clearly shows immune response in all participants and a correlation with age, gender and BMI: higher antibody titre was detected in younger people, in females and in normal-weight participants (BMI <25).

Our participants were grouped according to the BMI classification, which is based on the western classification of obesity(10). BMI is considered a crude measure that does not distinguish between under skin and visceral fat accumulation. Indeed, people in some areas of Asia, Middle East and Latin America tend to accumulate visceral fat at lower BMI. However, we believe that this classification could favor data comparison among different studies, as non-western researchers, like those in China, are also reporting their results using Western definitions of BMI(11).

The effectiveness of COVID-19 vaccines in people with obesity is a critical issue. Since obesity is a major risk factor for morbidity and mortality for patients with COVID-19(12), it is mandatory to plan an efficient vaccination program in this subgroup. Evidence suggests that SARS-CoV-2 infections are more severe and linger for about five days longer in people who are obese than in those who are lean(13).

The constant state of low-grade inflammation, present in overweight people, can weaken some immune responses, including those launched by T cells, which can directly kill infected cells(14).

Obesity is linked to less-diverse populations of microbes in the gut, nose and lung, with altered compositions and metabolic functions compared with those in lean individuals. Recently, researchers reported that changes of gut microbiome, by taking antibiotics, may alter responses to the flu vaccine(15). Moreover, vaccines against influenza, hepatitis B and rabies have shown reduced responses in those who are obese compared with those who are lean(16). To our knowledge, this study is the first to analyse COVID-19 vaccine response in correlation to BMI. Our data stresses the importance of close vaccination monitoring of obese people, considering the growing list of countries with obesity problems. According to the latest data from the World Health Organization, 39% of adults aged 18 years and over were overweight, and 13% were obese. If our data was to be confirmed by larger studies, giving obese people an extra dose of the vaccine or a higher dose could be options to be evaluated in this population.

Evidence from a recent meta-analysis suggest that COVID-19 exhibits differences in morbidity and mortality between sexes. Male patients have almost three times the odds of necessitating intensive treatment unit admission and higher odds of death compared to females(17). Our results also confirmed this difference in vaccine response. Women produce higher antibody titres in response to the trivalent inactivated seasonal influenza vaccination (TIV)(18, 19), as well as to most other pathogen vaccines(20). More specifically, females achieve equivalent protective antibody titres to males at half the dose of TIV(21), with serum testosterone levels inversely correlating with TIV antibody titres(22).

These findings imply that female, lean and young people have an increased capacity to mount humoral immune responses compared to male, overweight and older population. Although further studies are needed, this data may have important implications to the development of vaccination strategies for COVID-19, particularly in obese people. At the same time, we strongly believe that our results are extremely encouraging and useful for the scientific community.

All the authors have made substantial contributions to the work; all the authors approved the final manuscript. No honorarium grant or other form of payment was given to anyone of the authors to produce the manuscript. We have no conflict of interest to disclose.

## Data Availability

I confirm the availability of all data referred to in the manuscript

## REFERENCE

1. WHO, Draft Landscape of COVID-19 Candidate Vaccines,2021 (Accessed 14/02/ 2021 2021),https://www.who.int/publications/m/item/draft-landscape-of-covid-19-candidate-vaccines.

2. Polack FP, Thomas SJ, Kitchin N, Absalon J, Gurtman A, Lockhart S, et al. Safety and Efficacy of the BNT162b2 mRNA Covid-19 Vaccine. N Engl J Med. 2020;383(27):2603–15.

3. Verschoor CP, Singh P, Russell ML, Bowdish DM, Brewer A, Cyr L, et al. Microneutralization assay titres correlate with protection against seasonal influenza H1N1 and H3N2 in children. PLoS One. 2015;10(6):e0131531.

4. Chandrashekar A, Liu J, Martinot AJ, McMahan K, Mercado NB, Peter L, et al. SARS-CoV-2 infection protects against rechallenge in rhesus macaques. Science. 2020;369(6505):812–7.

5. Walsh EE, Frenck RW, Jr., Falsey AR, Kitchin N, Absalon J, Gurtman A, et al. Safety and Immunogenicity of Two RNA-Based Covid-19 Vaccine Candidates. N Engl J Med. 2020;383(25):2439–50.

6. Sahin U, Muik A, Vogler I, Derhovanessian E, Kranz LM, Vormehr Met al. BNT162b2 induces SARS-CoV-2 neutralising antibodies and T cells in humans. December 11, 2020 (https://www.medrxiv.org/content/10.1101/2020.12.09.20245175v1). preprint.

7. Krammer F, Srivasta K, the PARIS team, Simon V. Robust spike antibody responses and increased reactogenecity in seropositive individuals after a single dose of SARS-CoV-2 mRNA vaccine medRxiv 2021.02.29.21250653; doi: https://doi.org/10.1101/2021.01.29.21250653

8. Kulkarni PS, Hurwitz JL, Simoes EAF, Piedra PA. Establishing Correlates of Protection for Vaccine Development: Considerations for the Respiratory Syncytial Virus Vaccine Field. Viral Immunol. 2018;31(2):195–203.

9. Okba NMA, Muller MA, Li W, Wang C, GeurtsvanKessel CH, Corman VM, et al. Severe Acute Respiratory Syndrome Coronavirus 2-Specific Antibody Responses in Coronavirus Disease Patients. Emerg Infect Dis. 2020;26(7):1478–88.

10. Weir CB, Jan A. BMI Classification Percentile And Cut Off Points. [Updated 2020 Jul 10]. In:StatPearls [Internet]. Treasure Island (FL): StatPearls Publishing; 2020 Jan. Available from: https://www.ncbi.nlm.nih.gov/books/NBK541070/

11. Ledford H. How obesity could create problems for a COVID vaccine. Nature. 2020;586(7830):488–9.

12. Popkin BM, D. S, Green WD, Beck MA, Algaith T, Herbst CH, et al. Individuals with obesity and COVID-19: A global perspective on the epidemiology and biological relationships. Obes Rev. 2020;21(11):e13128.

13. Drucker DJ. Diabetes, obesity, metabolism, and SARS-CoV-2 infection: the end of the beginning. Cell Metab. 2021.

14. Vandanmagsar B, Youm YH, Ravussin A, Galgani JE, Stadler K, Mynatt RL, et al. The NLRP3 inflammasome instigates obesity-induced inflammation and insulin resistance. Nat Med. 2011;17(2):179–88.

15. Hagan T, Cortese M, Rouphael N, Boudreau C, Linde C, Maddur MS, et al. Antibiotics-Driven Gut Microbiome Perturbation Alters Immunity to Vaccines in Humans. Cell. 2019;178(6):1313–28 e13.

16. Frasca D, Blomberg BB. The Impact of Obesity and Metabolic Syndrome on Vaccination Success. Interdiscip Top Gerontol Geriatr. 2020;43:86–97.

17. Peckham H, de Gruijter NM, Raine C, Radziszewska A, Ciurtin C, Wedderburn LR, et al. Male sex identified by global COVID-19 meta-analysis as a risk factor for death and ITU admission. Nat Commun. 2020;11(1):6317.

18. Flanagan KL, Fink AL, Plebanski M, Klein SL. Sex and Gender Differences in the Outcomes of Vaccination over the Life Course. Annu Rev Cell Dev Biol. 2017;33:577–99.

19. Voigt EA, Ovsyannikova IG, Kennedy RB, Grill DE, Goergen KM, Schaid DJ, et al. Sex Differences in Older Adults’ Immune Responses to Seasonal Influenza Vaccination. Front Immunol. 2019;10:180.

20. Kleine P, Perthel M, Nygaard H, Hansen SB, Paulsen PK, Riis C, et al. Medtronic Hall versus St. Jude Medical mechanical aortic valve: downstream turbulences with respect to rotation in pigs. J Heart Valve Dis. 1998;7(5):548–55.

21. Engler RJ, Nelson MR, Klote MM, VanRaden MJ, Huang CY, Cox NJ, et al. Half-vs full-dose trivalent inactivated influenza vaccine (2004-2005): age, dose, and sex effects on immune responses. Arch Intern Med. 2008;168(22):2405–14.

22. Furman D, Hejblum BP, Simon N, Jojic V, Dekker CL, Thiebaut R, et al. Systems analysis of sex differences reveals an immunosuppressive role for testosterone in the response to influenza vaccination. Proc Natl Acad Sci U S A. 2014;111(2):869–74.

